# A Chatbot for the Management of Bipolar Disorder: Using Retrieval-Augmented Generation with an Open-Weight Large Language Model to Answer Clinical Questions Based on the CANMAT and ISBD 2018 Guidelines for Bipolar Disorder

**DOI:** 10.64898/2025.11.30.25341311

**Authors:** Yash Mali, Zejiao Zeng, Kayoung Heo, Grace Zhang, Antarip Kashyap, Jincheng Chen, Kamyar Keramatian, Gayatri Saraf, Marco Solmi, Edwin Tam, Sagar V. Parikh, Ayal Schaffer, Serge Beaulieu, Raymond Ng, Lakshmi N. Yatham, John-Jose Nunez

**Affiliations:** Department of Computer Science, University of British Columbia; Department of Psychiatry, University of British Columbia; Department of Linguistics, University of British Columbia; Department of Psychiatry, University of Ottawa; Department of Psychiatry, University of Michigan; Department of Psychiatry, University of Toronto; Department of Psychiatry, McGill University

## Abstract

**Objective:** Clinical practice guidelines support evidence-based care but are often underused due to complexity, time constraints, and navigation challenges. We investigated whether a conversational agent (chatbot) using an open-weight large language model (LLM) with retrieval-augmented generation (RAG) could provide guideline-consistent answers for bipolar disorder management based on the full 2018 CANMAT and ISBD guidelines, comparing against a system using only the base LLM.

**Method:** We developed a multi-step RAG-based chatbot that retrieves relevant guideline sections and generates responses using Llama 3.3 70B. Twenty-one clinical vignettes spanning all guideline sections were created. Six expert psychiatrists generated queries and were presented with paired responses without labels from two systems: one using the base Llama 3.3 70B model, the other RAG-enhanced. Responses rated guideline consistency on a three-point scale, and were analyzed using mixed-effects ordinal logistic regression.

**Results:** Experts evaluated 126 responses, of which 110 (87.3%) were rated as more or as correct as the baseline system. The RAG system produced 80 answers (63.5%) rated fully consistent with the guidelines versus 24 (19.0%) for baseline, and only 10 answers with major deviation (7.9%) versus 48 (38.1%) for baseline. Ordinal regression showed RAG responses were significantly more likely to be more correct (OR = 9.1, 95% CI 5.3–16.3, p < 0.001), which was consistent across all raters. Preference ratings favored RAG answers in 78.7% of cases. Performance varied by vignette, with some errors in both retrieval and reasoning.

**Conclusion:** The use of RAG with an open-weight model helped produce answers consistent with the CANMAT guidelines across vignettes that required adapting or combining guideline text, suggesting viability of a bipolar guideline chatbot. We identified areas to improve results and evaluation. Future work should explore additional retrieval strategies and LLMs, and test in more naturalistic settings.

## Introduction

Bipolar disorder is a prevalent and chronic psychiatric condition that presents a significant public health challenge globally ^1^, with a lifetime prevalence rate of bipolar I at 0.87% and bipolar II at 0.57% ^2,3^. Its management is complicated by the illness’s recurrent, multi-phased course, characterized by periods of wellness interrupted by episodes of mania, hypomania, and depression, along with its lifelong nature and high rates of comorbidities ^4^. Clinical guidelines, such as those developed by the Canadian Network for Mood and Anxiety Treatments (CANMAT) and the International Society for Bipolar Disorders (ISBD) ^5^, aim to enhance care through evidence-based recommendations. These guidelines used a rigorous overview of evidence, combined with expert clinical opinion, to produce pragmatic, hierarchical recommendations organized by treatment lines and levels-of-evidence. Despite the availability of clinical guidelines, their use in practice remains inconsistent ^6,7^, leading to suboptimal outcomes such as ineffective medications and inadequate consideration of comorbid conditions. Multiple barriers may play a role, including limited time and clinician skill ^8^, guideline complexity ^9^, challenges in applying recommendations to specific clinical scenarios ^10,11^, and difficulty navigating the documents ^12^.

To address these barriers, previous studies have examined digital tools such as C-IMPACT BD, a web application that provides rule-based recommendations from the CANMAT bipolar guidelines and has been shown to increase use of first-line therapies^13^. Recent advances in artificial intelligence offer additional opportunities to overcome barriers to guideline implementation. In particular, conversational agents, or “chatbots”, allow users to pose questions in natural language and receive support for treatment decisions^14^. These chatbots use large language models (LLMs) to answer questions. These models have been trained on language content from across the internet and may include knowledge relevant to clinical decision making^15^, though this information can be incomplete or incorrect, alongside more general sources such as internet forums. However, a challenge limiting their direct application in clinical settings is the generation of inaccuracies. These errors may arise from incorrect, biased, or outdated information, or as “hallucinations,” where the model fabricates content ^16^.

Retrieval-augmented generation (RAG)^17^ is a technique that can improve the accuracy of chatbot systems^18,19^. When a query is submitted, the RAG framework retrieves relevant information from domain-specific sources such as clinical guidelines, then passes the query, retrieved content, and system instructions to the LLM to generate a response. Initial studies have started to show that this technique can reduce hallucinations and improve factual relevance^20,21^. Examples of this approach in clinical settings are emerging, with LLMs being integrated with specialized texts and guidelines for managing specific conditions, such as thyroid disease^22^, liver disease^23,24^ and anticoagulation protocols for gastrointestinal procedures^25^. Because LLMs have a limited context window (the maximum text they can process at once), they cannot always include an entire guideline in a single prompt^26^. When context windows are smaller, RAG or a related technique is needed for retrieving relevant sections from large documents. Even with larger context windows, RAG helps focus responses on the most pertinent content rather than overwhelming the model with unnecessary text.

To our knowledge, RAG systems have not yet been studied for providing treatment recommendations in mental health using clinical guidelines. Existing applications have focused on other tasks, such as enhancing online mental health support system^27^, predicting the International Classification of Diseases (ICD) diagnostic code from psychiatry notes ^28^, and detecting depression ^29^. We identified one study that examined a chatbot for decision support in bipolar disorder treatment ^30^. This study did not use RAG; it augmented the chatbot’s prompt with an excerpt from the US Veterans Administration 2023 guidelines on bipolar depression^31^, and only tested recommendations for that mood state. Their augmented model prioritized expert-recommended medications 50.8% of the time, compared with 23.0% for the unaugmented model. The study used a proprietary, closed-weight model, GPT4-turbo, through OpenAI.

In this work, we investigate the use of RAG to provide treatment recommendations for bipolar disorder, covering all sections of the 2018 CANMAT guidelines, the world’s most cited for bipolar disorder. Our question set spans all mood states, both bipolar disorder types I and II, and addresses complex considerations such as age, comorbidities, side effects, special populations, and medication history. These vignettes often required reasoning that could not be satisfied by returning guideline text verbatim, as they involved synthesizing information from multiple sections and reconciling competing factors. We believe this reflects the complexity of treating mental illnesses like bipolar disorder and distinguishes our approach from prior guideline-based RAG systems, which have largely focused on simple knowledge retrieval ^24,33,34^. We also examined the use of an open-weight LLM, which offers advantages in privacy, interpretability, and reproducibility^32^. We hypothesized that our RAG system would outperform the base LLM for all expert raters, despite raters generating their own queries from vignettes, introducing variation in wording. This work aims to inform future applications of RAG systems in psychiatry and other areas of medicine where recommendations must integrate multiple data points from guidelines and patient-specific queries.

## Methods

### Vignette evaluation

We designed our evaluation based on recent assessments of clinical chatbots ^24,31–33^, asking expert raters to generate queries from clinical vignettes and score responses for consistency with the CANMAT 2018 guidelines. Six psychiatrists were initially recruited through the CANMAT network. All were guideline experts, either as co-authors or through regular clinical and educational use, and each had at least 10 years of experience treating bipolar disorder (range: 10–30 years). Raters received an orientation that included a walkthrough of a warm-up vignette, which was not scored. Due to an error, one participant did not receive most of the introduction and did not complete the warm-up case; this individual was excluded, and a seventh rater was subsequently recruited.

To avoid the risk of overfitting, where questions might be tailored to produce optimal answers for a fixed set of questions, we asked raters to write their own queries based on the provided vignettes rather than using the original wording. This approach also enabled evaluation of a more naturalistic and broader range of queries, as even minor differences in phrasing can significantly influence responses from systems using LLMs^34,35^.

To minimize courtesy bias, particularly because participants were recruited within the same network, raters were asked to blindly evaluate responses from two versions of the chatbot: one using RAG and one without, which we will refer to as the base model. We did not disclose how the two versions differed, only informing participants that they would be comparing two chatbot systems. The order in which responses appeared (left or right) was randomized for each vignette.

We created our primary scoring criterion by adapting a recently used scoring criterion^36^:

- **Correct:** Fully aligns with the 2018 CANMAT guidelines and is clinically actionable.
- **Inaccurate:** Largely correct but includes minor omissions or deviations that could lead to suboptimal decisions.
- **Wrong:** Contains major errors or contradictions that could compromise patient safety.

As a secondary outcome, we also asked participants to choose between the two responses by judging overall preference, considering readability, contextual applicability, and clarity in expressing complex clinical concepts, also based on a recent study^37^. The purpose of this was to provide an initial investigation as to whether a RAG system adversely affected these usability aspects.

### Clinical Vignettes Development

We developed 21 vignettes, adapting Perlis’ (2024)^31^ vignette format, to simulate clinical scenarios encountered by clinicians treating bipolar disorder, creating three vignettes from each of the seven sections of the 2018 CANMAT guidelines (excluding the introduction and conclusion). Vignettes are summarized in Table 1, with an example provided in Supplementary Table 1. To ensure clarity in age-related cases, older adult patients were assigned ages in the 70s, adolescents between 13 and 17 years, and all other cases between 20 and 59 years, with a mix of genders. The complexity of each vignette was intended to reflect questions that a community psychiatrist might reasonably pose in practice. Scenarios were designed to require integration of multiple guideline components (for example, combining recommendations for bipolar I maintenance with strategies to mitigate weight gain) or to adapt the guidelines to the clinical scenario, such as excluding previously failed medications. However, the degree of complexity and reasoning varied depending on how raters formulated their queries, particularly for cases 1 and 4.

**Table 1.**
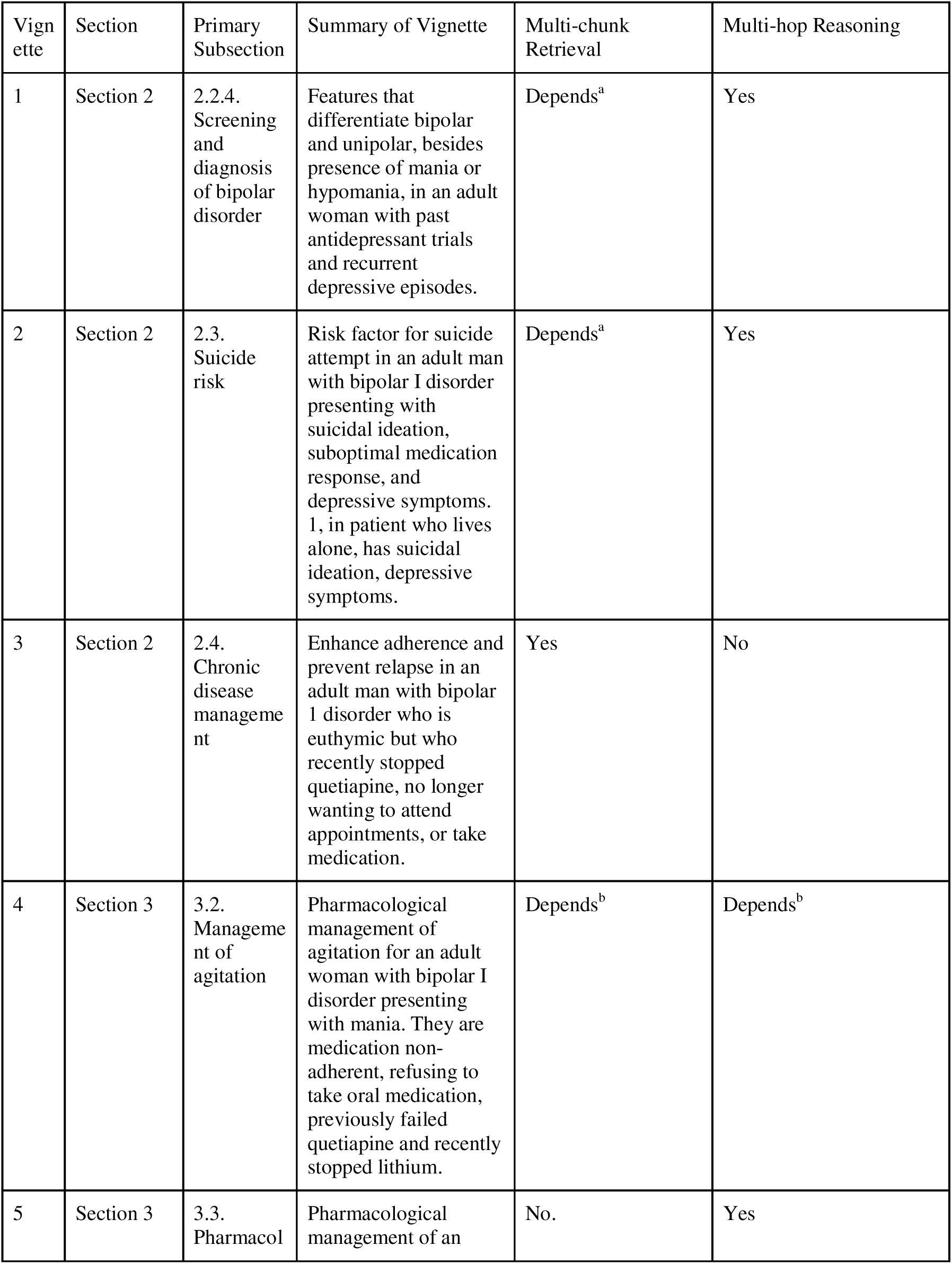
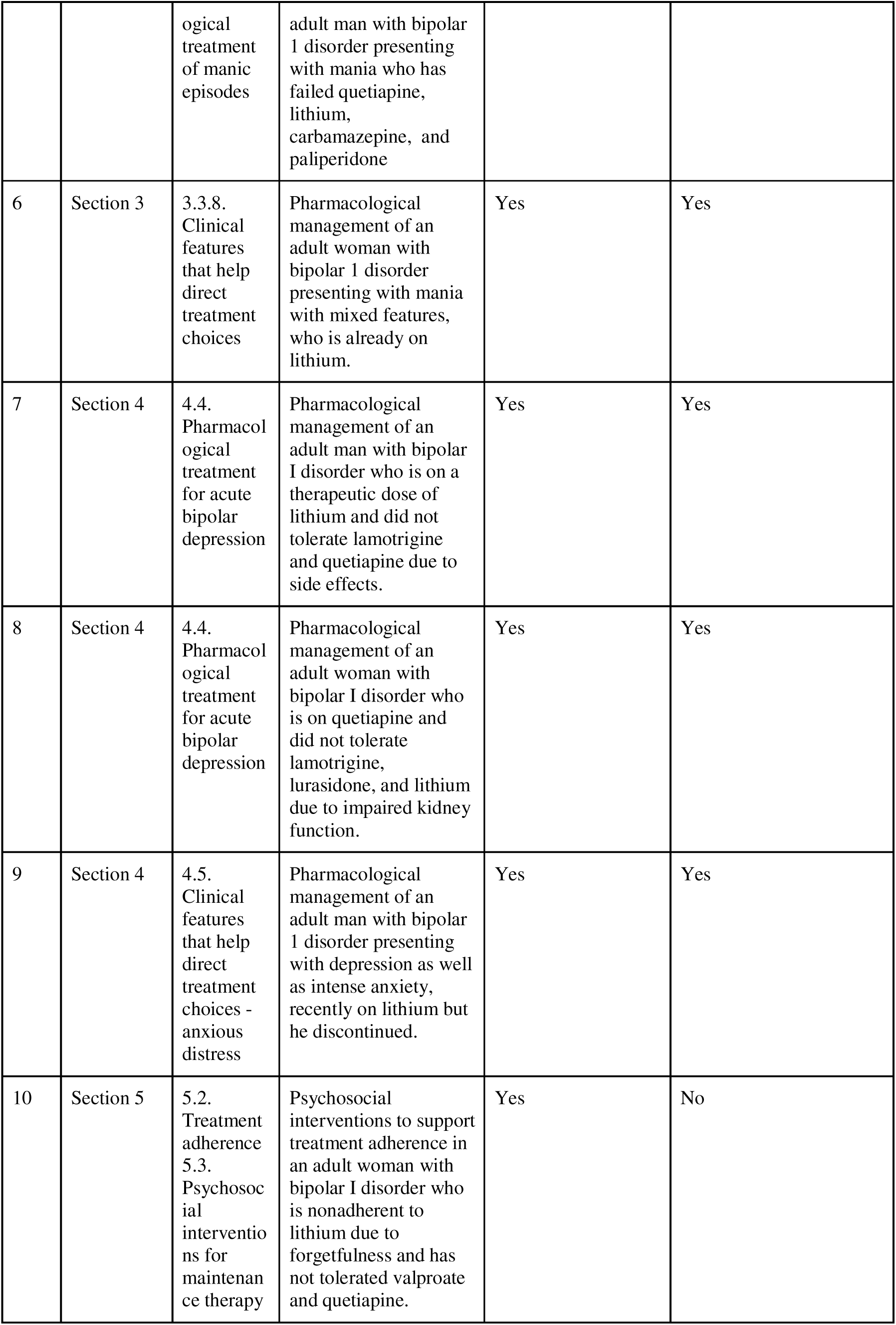
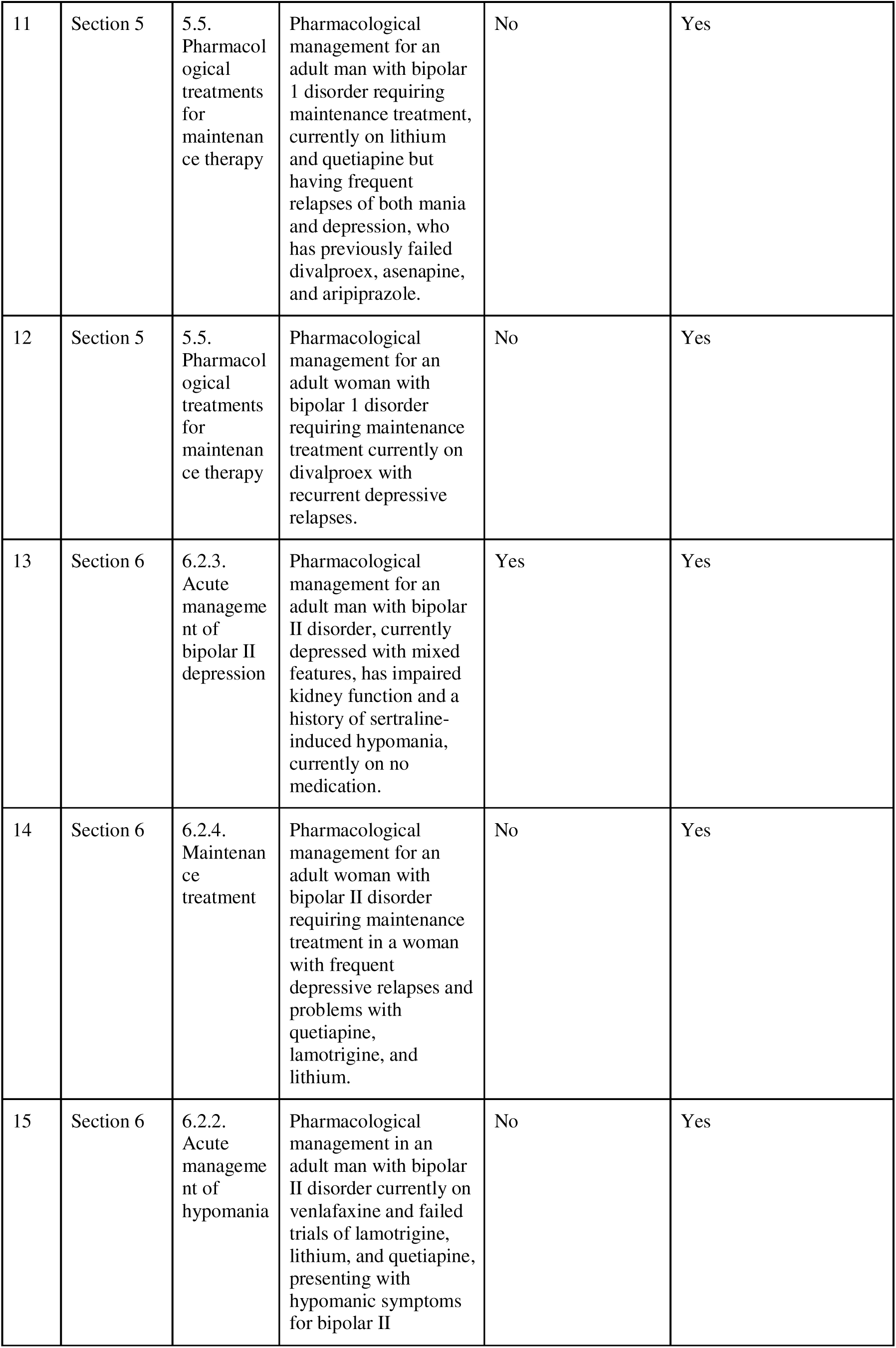
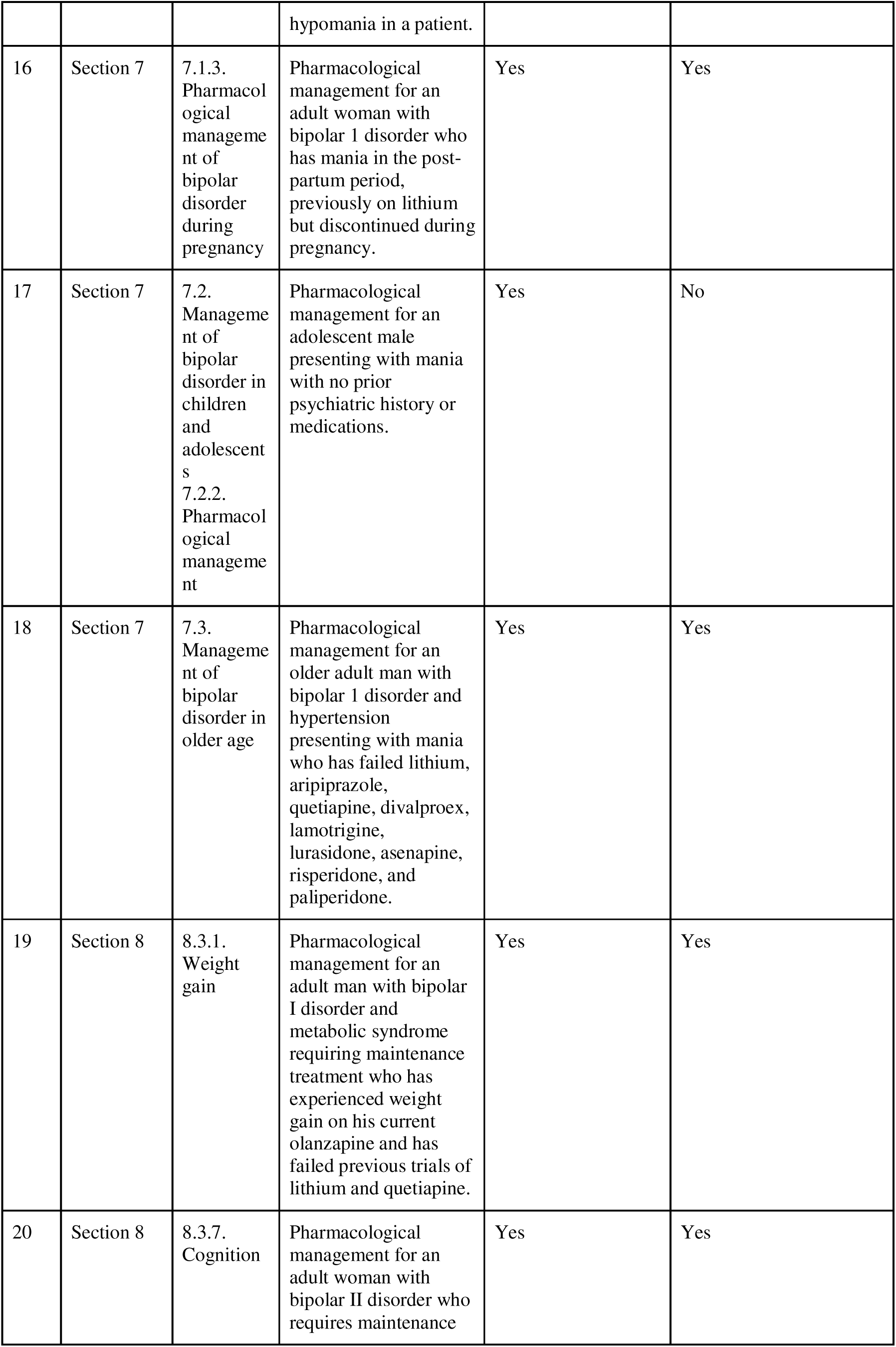
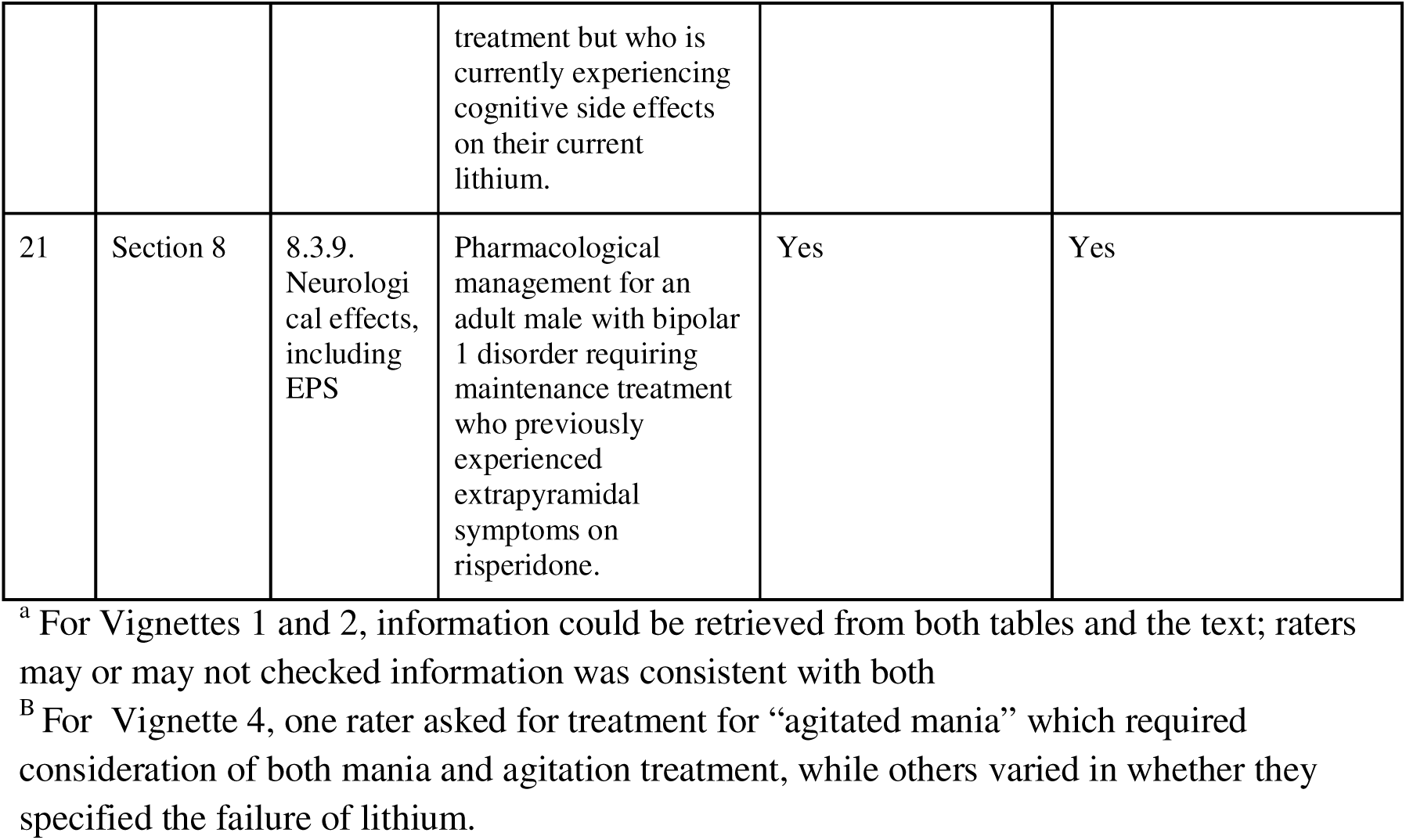
Clinical Vignettes. . “Multi-chunk” indicates answers requiring information from more than one part of the guidelines. “Multi-hop” refers to cases where non-trivial reasoning is needed, so responses cannot simply restate retrieved text verbatim.

### Chatbot Design

#### Document Preparation and Retrieval

We created a database of guideline content by scraping the CANMAT 2018 bipolar disorder website, converting paragraphs, tables, and figures into text. Each chunk was annotated with its section, subsection, and any referenced tables or figures to support downstream filtering.

Our system uses a multi-step RAG pipeline (Figure 1). First, user queries are classified into one of five categories: bipolar I mania, bipolar I depression, bipolar I maintenance, bipolar II, or general query. This classification, performed by a lightweight Mistral 7B model^38^, restricts the subsequent vector search to relevant guideline sections.

**Figure 1.**
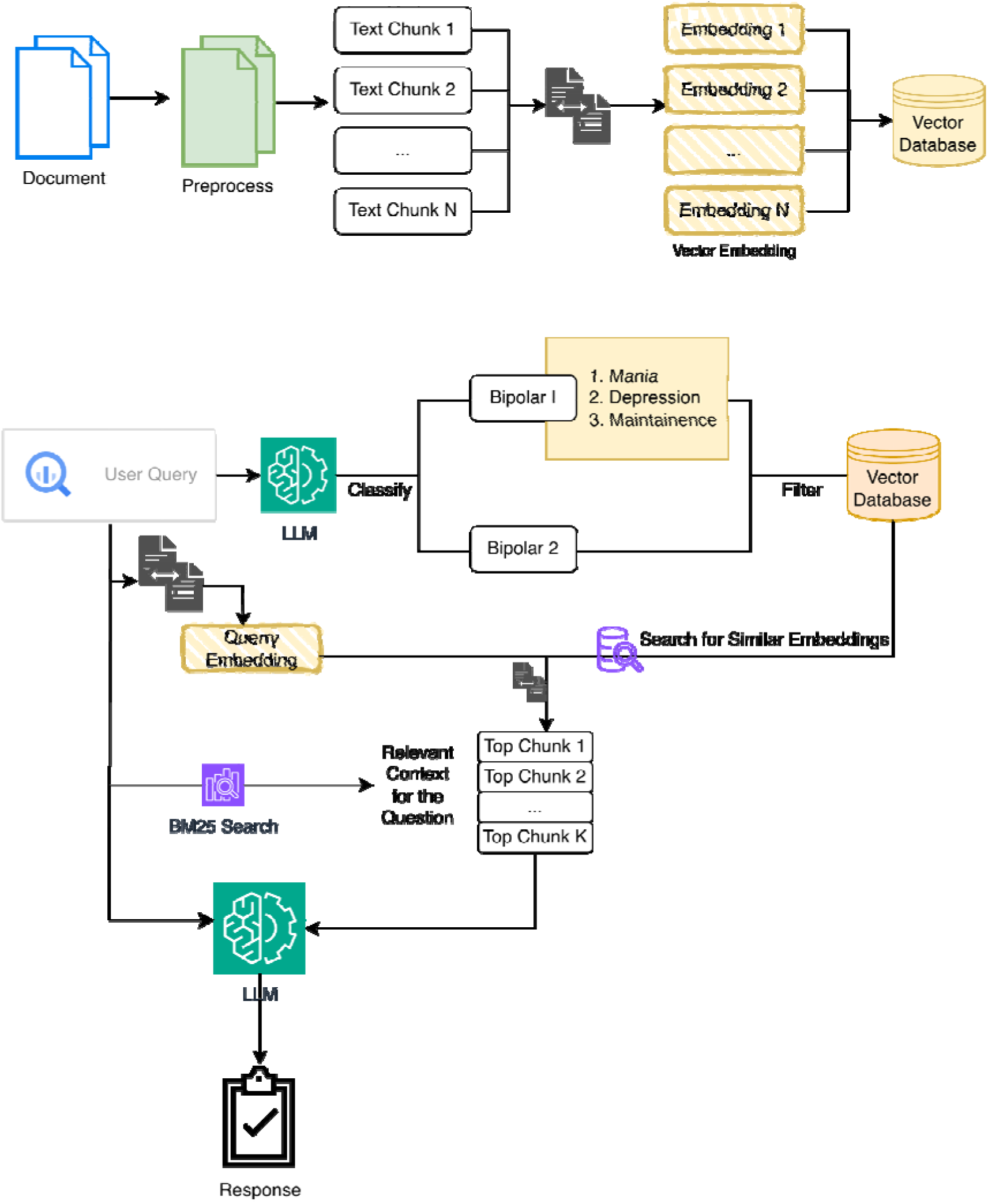
Diagram of our Multi-step Retrieval-augmented Generative System.

To improve retrieval accuracy, we implemented a hybrid approach that combines dense and sparse methods: embedding-based search (Qwen/Qwen3-Embedding-0.6B)^39^ for semantic similarity and BM25^40^ for exact keyword matching. Section headers are boosted during BM25 scoring. When a query includes header terms such as “cognition,” “treatment,” or “adverse effects,” scores for chunks in that section are multiplied by two. References to tables or figures in retrieved sections are appended to the context for final response generation.

#### Response Generation using a Large Language Model

For answer generation in our RAG system, we employed Llama 3.3 70B, which was the most capable open-weight model available at the study’s inception. Llama 3.3 70B has demonstrated performance comparable to leading commercial models released around the same time, such as GPT-4^41,42^ showing similar scores on benchmarks such as GPQA, Math 500 and BFC^43^, though results vary by task.

Prompt design was refined through collaboration between team members with expertise in computer science and psychiatry (Supplementary Table 2). The system prompt instructed the model to use only the retrieved context when reasoning and answering questions, reducing hallucinations. We used the Chain-of-Thought (CoT)^44^ technique in prompt engineering to improve LLM reasoning. The prompt guides the LLM to first gather patient attributes, such as disorder subtype, mood state, population, and then apply hierarchical reasoning to decide the treatment line needed, exclude medications based on comorbidities, side effects, evidence level, or user preferences, reflecting a clinician’s decision-making process.

For our evaluation, we maintained nearly identical prompts for the RAG and baseline versions to isolate the effect of retrieval augmentation; the only modification was removing instructions related to utilizing the retrieved context in the baseline model, as it operated without access to retrieved documents. The complete prompts are provided in the Supplementary Table 2.

### Cloud Deployment and User Interface

The user interface (UI) of the chatbot consists of a standard chat window with the CANMAT bipolar guidelines displayed alongside (Figure 2, a). Responses include the retrieved “chunks”, linking to the relevant subsections of the guidelines.

**Figure 2.a.**
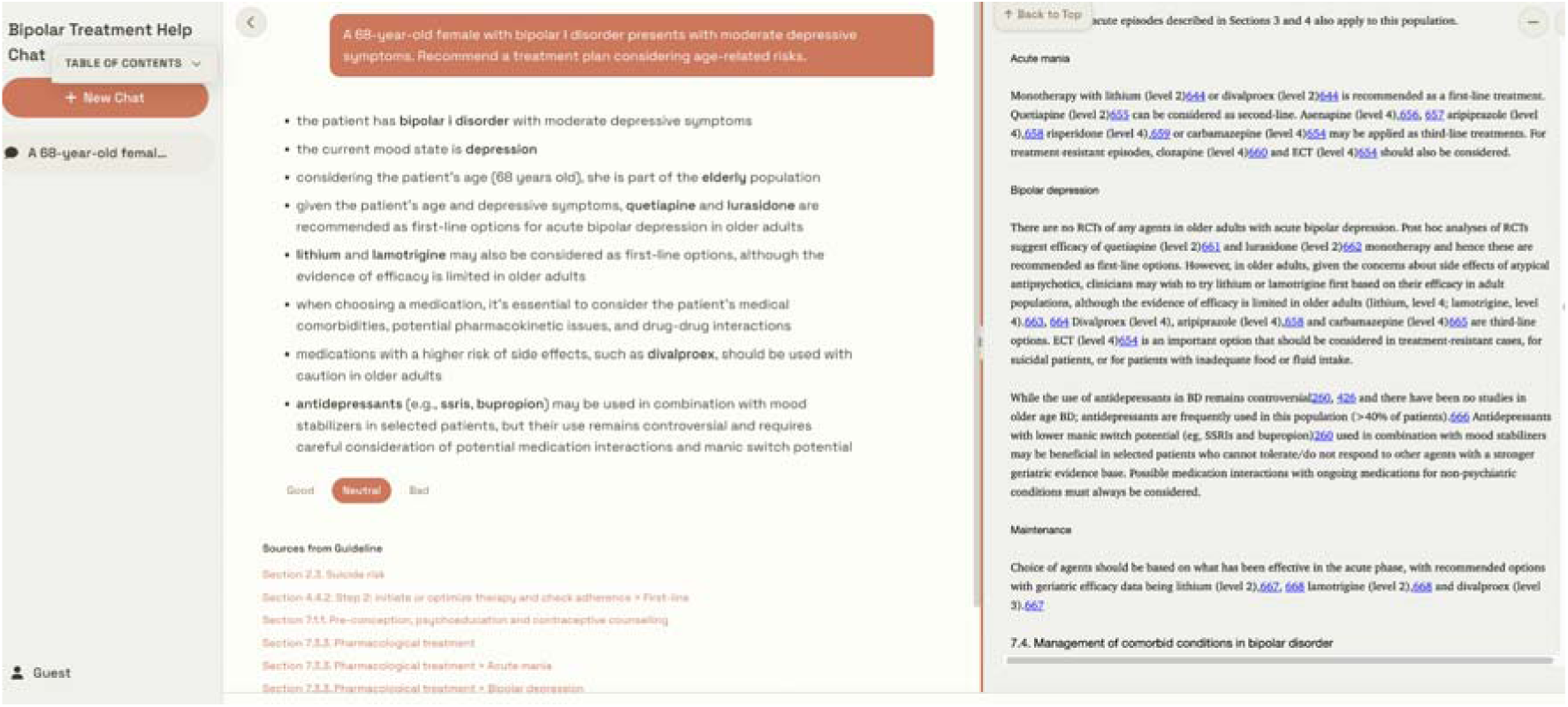
Our Chatbot in Standard Mode, Linking to Relevant Parts of the Guidelines Which Can be Viewed.

For the interface of the chatbot used in the evaluation, links to the subsections were not included so as not to differentiate the responses further from the base model. Our evaluation interface displays responses side by side and lists three rating options for each answer, as well as a button to choose the preferred response (Figure 2.b).

**Figure 2.b.**
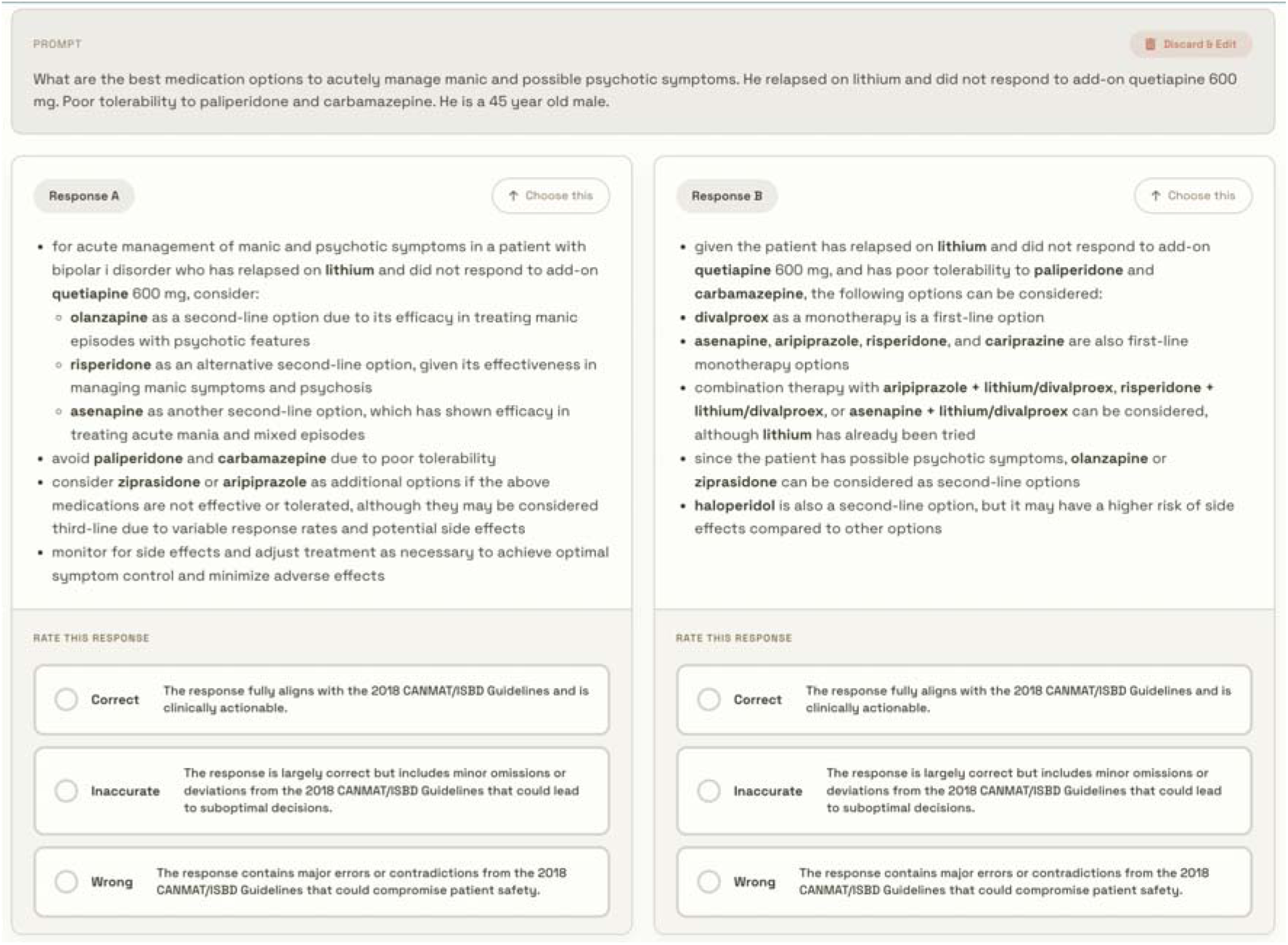
Our Chatbot as Seen During Our Evaluation.

We deployed our system on Amazon Web Services (AWS) through our institution’s AWS account. The chatbot itself is hosted on AWS Amplify, and it is built using Node.js. The vector search runs directly in the user’s web browser. All LLM inference tasks are handled by AWS Bedrock and embedding model inference by Sagemaker. Further architectural details are available on our GitHub repository.

### Statistical Analysis

We analyzed rating outcomes using a mixed-effects ordinal logistic regression model implemented in the R package *brms* ^45^. This approach was selected because the ratings (i.e., wrong, inaccurate, and correct) are inherently ordered, and the model accommodates this structure without collapsing categories into binary outcomes. Including random intercepts for participants and vignettes accounts for variability in rater tendencies and vignette difficulty, thereby improving the generalizability of the findings. The cumulative logit link estimates the effect of rating method (RAG vs baseline) on the odds of being in a higher category, under the proportional odds assumption. We summarized these predictions to characterize individual-level tendencies, providing mean posterior probabilities and 95% credible intervals for each rater under both conditions. This approach allows us to assess whether the observed group-level effect of RAG versus baseline is consistent across participants (Supplementary Document 1).

### Data Availability

The code used for our chatbot and evaluation will be available on a public GitHub repository upon publication. Given the potential for automatic scraping systems to use any publicly available data to train LLMs, which may impact planned immediate short-term work, the vignettes and question–answer pairs will be made available upon reasonable request to the corresponding author within two years of publication.

## Results

Our system performed better than the base model on the clinical vignettes. In total, raters evaluated 126 responses, 80 (63.5%) of them from the RAG system were marked as fully consistent (correct), compared to 24 (19%) from the base model. 36 RAG responses (28.6%) were marked as having minor deviations or omissions (inaccurate), compared to 54 (42.9%) from the base model. 10 RAG responses (7.9%) were marked as having major deviations (wrong), compared to 48 (38.1%) from the base model (Supplementary Table 3 and Supplementary Figure 1). The RAG system provided answers that were statistically significantly more likely to be more correct than the baseline model (OR = 9.1, 95% CI 5.3– 16.3, p < 0.001). Across 84 ratings (66.7%), RAG provided an answer that was more correct than the base model, 26 (20.6%) had the same level of rating, and 16 (12.7%) less correct. In total, 110 (87.3%) of the RAG responses were rated as more or as correct as the baseline system. Mixed-effects ordinal logistic regression confirmed this advantage persisted across all individual raters, each showing posterior probabilities of superiority equal to 1.0 for every rater.

For our secondary analysis, overall, raters preferred the RAG response in 96 (78.7%) ratings out of 122 preference ratings. Raters did not provide preferences for four ratings. In the 26 ratings where the experts rated both answers as the same level of correctness, raters preferred 18 (69.2%) of responses from RAG. In the 16 ratings where the baseline was correct, the baseline responses were preferred.

For the RAG system, performance varied across vignettes. Some cases, such as Vignettes 1 and 4, had all six responses rated correct, whereas Vignette 9 had none marked correct and five rated inaccurate, with the base model performing better by producing two correct answers (Table 2). Vignettes 15 and 18 showed mixed ratings, with three responses marked correct, two inaccurate, and one wrong. In several cases (e.g., Vignettes 2, 3, and 19), both models achieved the same number of correct responses.

**Table 2.**
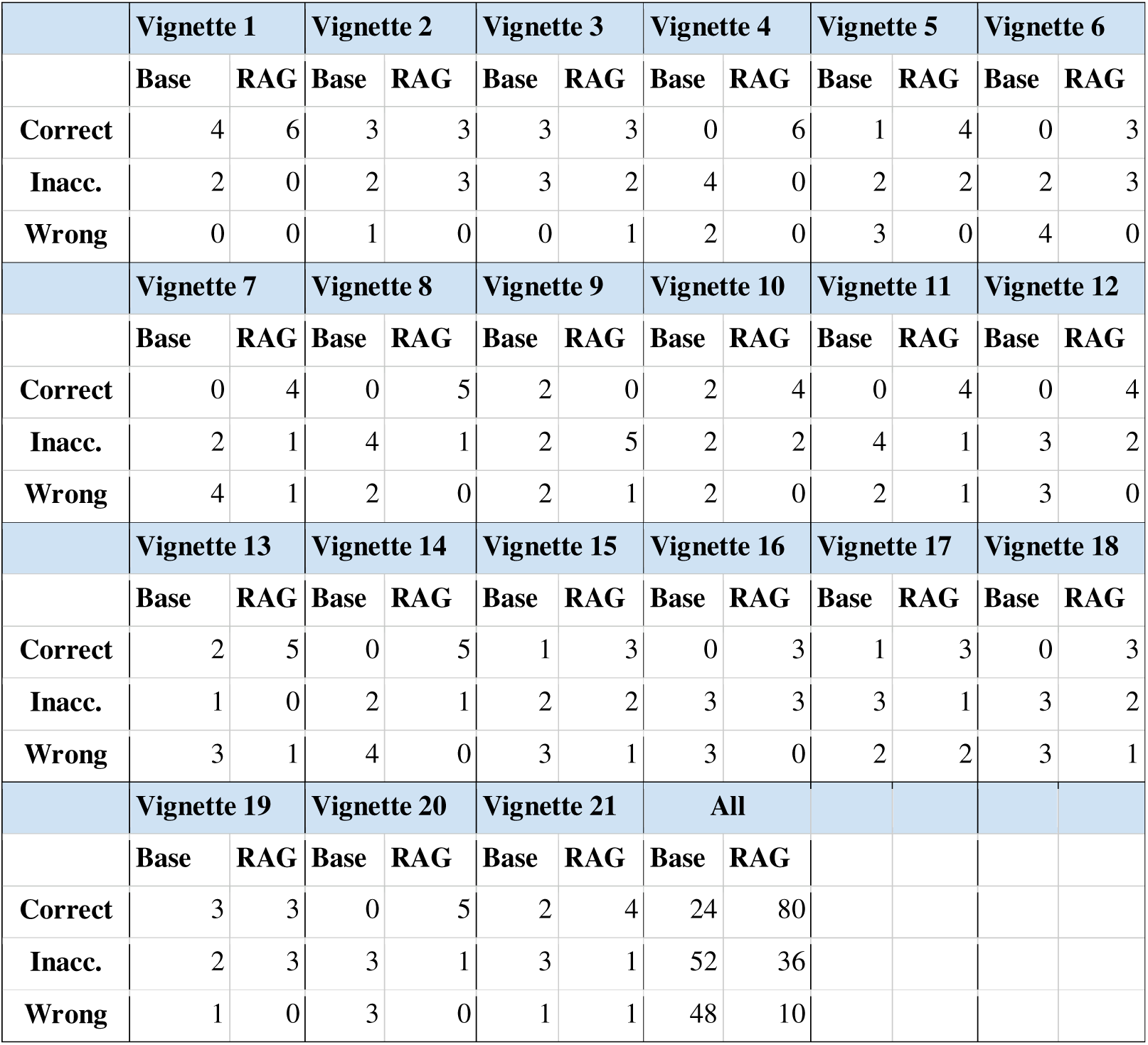
Chatbot Ratings for Each Vignette.

We also observed variability among raters in how many responses they marked as correct. The strictest rater judged only eight RAG responses as correct and marked six as wrong, whereas the most lenient rater marked 18 responses as correct and none as wrong (Figure 3).

**Figure 3.**
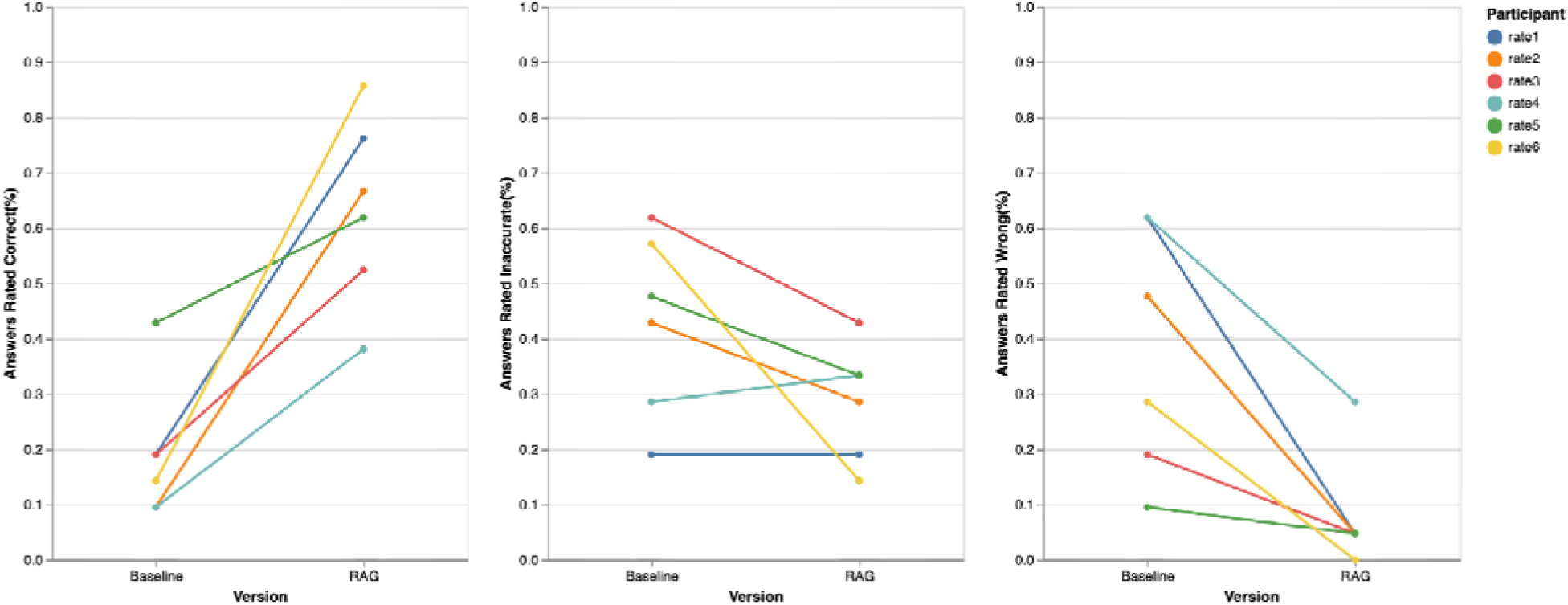
Comparison of ratings across six expert raters between the baseline and retrieval-augmented generation (RAG) responses.

Of the 10 RAG-generated responses rated as wrong, six appeared to result from inadequate retrieval context. Two of these errors occurred in Vignette 17, which involved managing mania in an adolescent patient, while the remaining errors were distributed across other vignettes.

## Discussion

This is the first study to evaluate the use of RAG for generating treatment recommendations for bipolar disorder based on the 2018 CANMAT guidelines. RAG combines a large language model with retrieval of relevant guideline content to support answers that require synthesizing information rather than simple text lookup. Our goal was to design a system that could provide just-in-time answers to clinical questions across all mood states, bipolar disorder subtypes, and comorbidities, reflecting the complexity of real-world decision-making. Our findings indicate that the multi-step RAG system, built on open-weight models, produces answers that align with CANMAT guidelines at a substantially higher rate than the base model without RAG, which achieved only 19.1% correct responses and 38.1% with major errors. In contrast, RAG yielded 63.5% fully guideline-consistent responses and only 7% with major errors, demonstrating numerically better performance than a prior study assessing treatment decisions from an augmented GPT-4 Turbo model for bipolar depression alone^30^, though differences in study design limit direct comparison. We believe our study provides an initial demonstration of how RAG can be used with clinical guidelines in psychiatry for scenarios involving multiple considerations, such as age, previous medication responses, special populations, and comorbidities, providing directions for further improvement when seeking to design systems to help in complex clinical scenarios.

Our results provide an example of how RAG can support mental health treatment recommendations. The best performing vignette, Vignette 4, had all responses marked correct, likely because it required straightforward reasoning; the answer was simply the first-line treatment options for agitation without modification. However, one rater’s query illustrated how phrasing can vary by asking for treatment for “agitated mania” rather than just agitation. The system interpreted this composite query and returned preferred treatments for both mania and agitation, demonstrating its flexibility in handling nuanced language. Other high-performing vignettes, such as Vignette 8 and Vignette 20, demonstrated the system’s ability to generate recommendations based on scenario-specific factors. The former required adapting guideline tables on bipolar I depression treatment after excluding multiple failed medications, while the latter required integrating content from multiple sections discussing maintenance treatment for bipolar II and strategies to reduce cognitive symptoms. One rater judged the RAG system’s performance as particularly strong, marking 85.7% of its responses as correct and the remainder as inaccurate, which underscores the potential of this approach, although outcomes may depend on query formulation and interpretation.

Evaluation of the RAG-based chatbot revealed three key challenges: variability in retrieval accuracy, limitations in multi-step reasoning, and inconsistencies between expert scoring and the system’s design objectives. While most RAG responses were correct, the 7% with major errors seemed to have been related to retrieval issues, particularly in Vignette 17. This case required recommendations specific to the subsection on treating adolescents and youths. Despite the explicit instruction in the prompt for LLM to consider age factors, the retrieval component often failed to interpret numeric ages as indicators of youth, instead relying on explicit keywords such as “youth” or “adolescent.” When these were absent, the model defaulted to adult-specific recommendations. A substantial portion of errors appeared related to LLM weaknesses in multi-step reasoning. For example, in Vignette 9, the RAG chatbot appeared to struggle with balancing information retrieved from chunks on acute bipolar I depression, anxious distress, and comorbid anxiety. This sometimes led to incomplete responses, such as failing to recommend olanzapine–fluoxetine even though it is listed in the comorbid-anxiety subsection, likely because the acute-depression table designates it as a second-line option. In contrast, the baseline model, which was not constrained by specific retrieved chunks, recommended this combination more consistently, contributing to its superior performance on this vignette. Additionally, some responses were rated inaccurate or wrong because experts applied different rating standards or clinical judgment beyond the guideline scope, despite instructions to evaluate solely on consistency with the guideline. For instance, in Vignette 17, identical responses from the baseline model were rated “correct” by one expert and “inaccurate” by another, underscoring variability in scoring. Similarly, a rater judged a RAG response to Vignette 5 (Supplementary Table 1) as inaccurate because it suggested risperidone after paliperidone failure, noting that paliperidone is the active metabolite of risperidone—a clinically reasonable interpretation but not specified in the guidelines. This underscores a misalignment between system design and evaluation criteria: the chatbot was built for document-grounded reasoning, whereas experts often applied a physician standard, contributing to lower-rated accuracy. Future improvements could include integrating external clinical knowledge bases to better emulate clinical reasoning.

Future improvements should focus on two key limitations that can sometimes lead to errors in RAG systems: incomplete retrieval of relevant guideline sections and challenges with multi-step reasoning. Advanced RAG techniques and newer models could help address these issues. Our current search approach retrieves information in a single step, which makes it difficult to answer questions that require combining details from different sections of the guidelines.^46^ Approaches such as PlanRAG^47^ and self-correction^48^ mechanisms, along with retrieval-enhanced embedding models, warrant exploration. To improve reasoning, two directions appear promising. First, testing newer LLMs, including open-weight models such as gpt-oss^49^ and state-of-the-art commercial systems Gemini 3^50^, combined with a structured clinical reasoning framework for prompt design, could enhance performance. Second, hybrid architectures that integrate rule-based components with LLMs may offer greater reliability.

This strategy aligns with existing tools such as the C-IMPACT web application ^51^, which provides rule-based recommendations for the CANMAT bipolar guidelines. Although C-IMPACT does not currently cover all scenarios tested in this study, it could be expanded. However, the upcoming update to the CANMAT bipolar guidelines would require ongoing rule development, highlighting the adaptability advantage of systems that minimize manual rule creation.

Further evaluation will be needed to test iterative improvement. Our methodology highlighted challenges related to rater availability and variability. Most raters required more than two hours to complete 21 ratings, making it impractical to compare different system outcomes and continuous optimization. Future studies should explore ways to provide robust evaluations while reducing the time burden on guideline experts, who are often clinician-scientists with limited availability. One option is to develop marking rubrics for vignettes, allowing less experienced raters, such as psychiatry residents, to complete evaluations with an expert available to resolve disagreements. Rubrics could also support automated evaluation, although human oversight would likely still be necessary. To avoid overfitting, rubrics should accommodate variability in queries and could be created after external clinicians generate the queries rather than the chatbot development team.

While we have discussed various limitations above, such as testing only one, open-weight model and variability in our evaluations, other limitations should be considered. Our study did not evaluate the impact of gender or other equity considerations on responses. While we believe our study was based on scenarios with more complexity than many others in RAG studies so far, they still represent simplifications compared to real-world patients, and our evaluation solely evaluated single-turn question answering, deferring the evaluation of real-world, dynamic multi-turn conversations to future work.

Despite these limitations, our study provides a foundation for further use of RAG chatbots to support decision-making in mental health, while highlighting multiple directions for improvement. Although our results showed that responses still contained minor and occasional major deviations from the guidelines, future work could examine how a chatbot with this level of performance functions in more naturalistic settings, such as with clinicians at varying career stages or trainees who lack guideline expertise. A prior study in bipolar depression found that community-based clinicians selected guideline-consistent answers for only 23.1% of vignettes, whereas a simple prompt-augmented chatbot using GPT 4 Turbo achieved 50.8% ^30^. This suggests that even imperfect systems may meaningfully improve care. In the future, explainable AI^52^ could enhance such tools by highlighting relevant guideline text and reasoning, enabling error checking and supporting clinician education. Ultimately, we believe this work advances efforts to integrate AI into clinical workflows, supporting evidence-based care for individuals with bipolar disorder and other conditions.

## Acknowledgements

Technical guidance, project coordination, and computing resources were provided by the University of British Columbia Cloud Innovation Centre.

## In Supplementary Materials

● Supplementary Table 1: Example of a Clinical Vignette
● Supplementary Table 2. Prompt for baseline Llama Model and RAG-based Llama Model
● Supplementary Table 3. Comparing the Rating of RAG with the Base Model
● Supplementary Figure 1: Confucian Matrix
● Supplementary Document 1: Ordinal Mixed-Effects Model: Summary and Posterior Analysis

## References

1. Singh B, Swartz HA, Cuellar-Barboza AB, et al. Bipolar disorder. The Lancet. 2025;406(10506):963–978. doi:10.1016/S0140-6736(25)01140-7

2. McDonald KC, Bulloch AGM, Duffy A, et al. Prevalence of Bipolar I and II Disorder in Canada. Can J Psychiatry. 2015;60(3):151–156. doi:10.1177/070674371506000310

3. Schaffer A, Cairney J, Cheung A, Veldhuizen S, Levitt A. Community Survey of Bipolar Disorder in Canada: Lifetime Prevalence and Illness Characteristics. Can J Psychiatry. 2006;51(1):9–16. doi:10.1177/070674370605100104

4. Fountoulakis KN, Vieta E, Siamouli M, et al. Treatment of bipolar disorder: a complex treatment for a multi-faceted disorder. Ann Gen Psychiatry. 2007;6(1):27. doi:10.1186/1744-859X-6-27

5. Yatham LN, Kennedy SH, Parikh SV, et al. Canadian Network for Mood and Anxiety Treatments (CANMAT) and International Society for Bipolar Disorders (ISBD) 2018 guidelines for the management of patients with bipolar disorder. Bipolar Disord. 2018;20(2):97–170. doi:10.1111/bdi.12609

6. Baek JH, Ha K, Yatham LN, et al. Pattern of Pharmacotherapy by Episode Types for Patients With Bipolar Disorders and Its Concordance With Treatment Guidelines. Journal of Clinical Psychopharmacology. 2014;34(5):577. doi:10.1097/JCP.0000000000000175

7. Paterniti S, Bisserbe JC. Pharmacotherapy for bipolar disorder and concordance with treatment guidelines: survey of a general population sample referred to a tertiary care service. BMC Psychiatry. 2013;13(1):211. doi:10.1186/1471-244X-13-211

8. Heselmans A, Donceel P, Aertgeerts B, Van De Velde S, Ramaekers D. The attitude of Belgian social insurance physicians towards evidence-based practice and clinical practice guidelines. BMC Fam Pract. 2009;10(1):64. doi:10.1186/1471-2296-10-64

9. Wolf JS, Hubbard H, Faraday MM, Forrest JB. Clinical practice guidelines to inform evidence-based clinical practice. World J Urol. 2011;29(3):303–309. doi:10.1007/s00345-011-0656-5

10. Mansfield CD. Attitudes and behaviors towards clinical guidelines: the clinicians’ perspective. BMJ Quality & Safety. 1995;4(4):250–255. doi:10.1136/qshc.4.4.250

11. Carlsen B, Bringedal B. Attitudes to clinical guidelines--do GPs differ from other medical doctors? BMJ Qual Saf. 2011;20(2):158–162. doi:10.1136/bmjqs.2009.034249

12. Foy MA. Clinical guidelines: Must we follow them? Bone & Joint 360. 2016;5(4):42–43. doi:10.1302/2048-0105.54.360450

13. Kozicky JM, Schaffer A, Beaulieu S, McIntosh D, Yatham LN. Use of a point-of-care web-based application to enhance adherence to the CANMAT and ISBD 2018 guidelines for the management of bipolar disorder. Bipolar Disord. 2022;24(4):392–399. doi:10.1111/bdi.13136

14. Liu S, Wright AP, Patterson BL, et al. Using AI-generated suggestions from ChatGPT to optimize clinical decision support. J Am Med Inform Assoc. 2023;30(7):1237–1245. doi:10.1093/jamia/ocad072

15. Kung TH, Cheatham M, Medenilla A, et al. Performance of ChatGPT on USMLE: Potential for AI-assisted medical education using large language models. PLOS Digit Health. 2023;2(2):e0000198. doi:10.1371/journal.pdig.0000198

16. Augenstein I, Baldwin T, Cha M, et al. Factuality Challenges in the Era of Large Language Models. *arXiv*. Preprint posted online October 11, 2023. doi:10.48550/arXiv.2310.05189

17. Lewis P, Perez E, Piktus A, et al. Retrieval-Augmented Generation for Knowledge-Intensive NLP Tasks. *arXiv*. Preprint posted online April 13, 2021. doi:10.48550/arXiv.2005.11401

18. Yu H, Gan A, Zhang K, Tong S, Liu Q, Liu Z. Evaluation of Retrieval-Augmented Generation: A Survey. In: Vol 2301. 2025:102–120. doi:10.1007/978-981-96-1024-2_8

19. Retrieval-Augmented Generation for Large Language Models: A Survey. Accessed November 13, 2025. https://arxiv.org/html/2312.10997v5

20. Liu S, McCoy AB, Wright A. Improving large language model applications in biomedicine with retrieval-augmented generation: a systematic review, meta-analysis, and clinical development guidelines. J Am Med Inform Assoc. 2025;32(4):605–615. doi:10.1093/jamia/ocaf008

21. Woo JJ, Yang AJ, Olsen RJ, et al. Custom Large Language Models Improve Accuracy: Comparing Retrieval Augmented Generation and Artificial Intelligence Agents to Noncustom Models for Evidence-Based Medicine. Arthroscopy. 2025;41(3):565–573.e6. doi:10.1016/j.arthro.2024.10.042

22. Shin M, Song J, Kim MG, Yu HW, Choe EK, Chai YJ. Thyro-GenAI: A Chatbot Using Retrieval-Augmented Generative Models for Personalized Thyroid Disease Management. J Clin Med. 2025;14(7):2450. doi:10.3390/jcm14072450

23. Ge J, Sun S, Owens J, et al. Development of a liver disease-specific large language model chat interface using retrieval-augmented generation. Hepatology. 2024;80(5):1158–1168. doi:10.1097/HEP.0000000000000834

24. Kresevic S, Giuffrè M, Ajcevic M, Accardo A, Crocè LS, Shung DL. Optimization of Hepatological Clinical Guidelines Interpretation by Large Language Models: A Retrieval Augmented Generation-Based Framework. npj Digit Med. 2024;7(1):1–9. doi:10.1038/s41746-024-01091-y

25. Malik S, Kharel H, Dahiya DS, et al. Assessing ChatGPT4 with and without retrieval-augmented generation in anticoagulation management for gastrointestinal procedures. Ann Gastroenterol. 2024;37(5):514–526. doi:10.20524/aog.2024.0907

26. Xu P, Ping W, Wu X, et al. Retrieval meets Long Context Large Language Models. *arXiv*. Preprint posted online January 23, 2024. doi:10.48550/arXiv.2310.03025

27. Yang D, Zhu J, Wu H, Tan M, Li C, Yang M. CascadeRCG: Retrieval-Augmented Generation for Enhancing Professionalism and Knowledgeability in Online Mental Health Support. In: Companion Proceedings of the ACM on Web Conference 2025. WWW ’25. Association for Computing Machinery; 2025:1465–1469. doi:10.1145/3701716.3715466

28. Boggavarapu L, Srivastava V, Varanasi AM, Lu Y, Bhaumik R. Evaluating Enhanced LLMs for Precise Mental Health Diagnosis from Clinical Notes. medRxiv. Preprint posted online March 10, 2025:2024.12.16.24317648. doi:10.1101/2024.12.16.24317648

29. Hou R, Teng S, Liu J, et al. Retrieval-Augmented Multimodal Depression Detection. *arXiv*. Preprint posted online October 29, 2025. doi:10.48550/arXiv.2511.01892

30. Perlis RH, Goldberg JF, Ostacher MJ, Schneck CD. Clinical decision support for bipolar depression using large language models. Neuropsychopharmacol. 2024;49(9):1412–1416. doi:10.1038/s41386-024-01841-2

31. Perlis RH, Goldberg JF, Ostacher MJ, Schneck CD. Clinical decision support for bipolar depression using large language models. Neuropsychopharmacology. 2024;49(9):1412–1416. doi:10.1038/s41386-024-01841-2

32. Masanneck L, Meuth SG, Pawlitzki M. Evaluating base and retrieval augmented LLMs with document or online support for evidence based neurology. npj Digit Med. 2025;8(1):137. doi:10.1038/s41746-025-01536-y

33. Wada A, Tanaka Y, Nishizawa M, et al. Retrieval-augmented generation elevates local LLM quality in radiology contrast media consultation. npj Digit Med. 2025;8(1):395. doi:10.1038/s41746-025-01802-z

34. Hond A de, Leeuwenberg T, Bartels R, et al. From text to treatment: the crucial role of validation for generative large language models in health care. The Lancet Digital Health. 2024;6(7):e441–e443. doi:10.1016/S2589-7500(24)00111-0

35. Rahmani HA, Siro C, Aliannejadi M, et al. Report on the 1st Workshop on Large Language Model for Evaluation in Information Retrieval (LLM4Eval 2024) at SIGIR 2024. *arXiv*. Preprint posted online August 9, 2024. doi:10.48550/arXiv.2408.05388

36. Masanneck L, Meuth SG, Pawlitzki M. Evaluating base and retrieval augmented LLMs with document or online support for evidence based neurology. npj Digit Med. 2025;8(1):137. doi:10.1038/s41746-025-01536-y

37. Zakka C, Shad R, Chaurasia A, et al. Almanac — Retrieval-Augmented Language Models for Clinical Medicine. NEJM AI. 2024;1(2). doi:10.1056/AIoa2300068

38. Jiang AQ, Sablayrolles A, Mensch A, et al. Mistral 7B. *arXiv*. Preprint posted online October 10, 2023. doi:10.48550/arXiv.2310.06825

39. Yang A, Li A, Yang B, et al. Qwen3 Technical Report. *arXiv*. Preprint posted online May 14, 2025. doi:10.48550/arXiv.2505.09388

40. Robertson S, Zaragoza H. The Probabilistic Relevance Framework: BM25 and Beyond. Found Trends Inf Retr. 2009;3(4):333–389. doi:10.1561/1500000019

41. Zhong W, Liu Y, Liu Y, et al. Performance of ChatGPT-4o and Four Open-Source Large Language Models in Generating Diagnoses Based on China’s Rare Disease Catalog: Comparative Study. Journal of Medical Internet Research. 2025;27(1):e69929. doi:10.2196/69929

42. Perlis R, Collins N. Can Open-Source AI Models Diagnose Complex Cases as Well as GPT-4? JAMA. 2025;333(17):1473–1475. doi:10.1001/jama.2025.2806

43. LLM Leaderboard 2025. Accessed November 15, 2025. https://www.vellum.ai/llm-leaderboard

44. Wei J, Wang X, Schuurmans D, et al. Chain-of-Thought Prompting Elicits Reasoning in Large Language Models. *arXiv*. Preprint posted online January 10, 2023. doi:10.48550/arXiv.2201.11903

45. Bürkner PC. brms: An R Package for Bayesian Multilevel Models Using Stan. Journal of Statistical Software. 2017;80:1–28. doi:10.18637/jss.v080.i01

46. Jiang Z, Sun M, Liang L, Zhang Z. Retrieve, Summarize, Plan: Advancing Multi-hop Question Answering with an Iterative Approach. arXiv. Preprint posted online January 30, 2025. doi:10.48550/arXiv.2407.13101

47. Lee M, An S, Kim MS. PlanRAG: A Plan-then-Retrieval Augmented Generation for Generative Large Language Models as Decision Makers. In: Duh K, Gomez H, Bethard S, eds. Proceedings of the 2024 Conference of the North American Chapter of the Association for Computational Linguistics: Human Language Technologies (Volume 1: Long Papers). Association for Computational Linguistics; 2024:6537–6555. doi:10.18653/v1/2024.naacl-long.364

48. Asai A, Wu Z, Wang Y, Sil A, Hajishirzi H. Self-RAG: Learning to Retrieve, Generate, and Critique through Self-Reflection. *arXiv*. Preprint posted online October 17, 2023. doi:10.48550/arXiv.2310.11511

49. OpenAI, Agarwal S, Ahmad L, et al. gpt-oss-120b & gpt-oss-20b Model Card. arXiv. Preprint posted online August 8, 2025. doi:10.48550/arXiv.2508.10925

50. Introducing Gemini 3: our most intelligent model that helps you bring any idea to life. https://blog.google/products/gemini/gemini-3/#gemini-3

51. Kozicky JM, Schaffer A, Beaulieu S, McIntosh D, Yatham LN. Use of a point-of-care web-based application to enhance adherence to the CANMAT and ISBD 2018 guidelines for the management of bipolar disorder. Bipolar Disorders. 2022;24(4):392–399. doi:10.1111/bdi.13136

52. Rizzo M, Veneri A, Albarelli A, Lucchese C, Nobile M, Conati C. A Theoretical Framework for AI Models Explainability with Application in Biomedicine. In: 2023 IEEE Conference on Computational Intelligence in Bioinformatics and Computational Biology (CIBCB). 2023:1–9. doi:10.1109/CIBCB56990.2023.10264877

