## Supplementary Material for "A Chatbot for the Management of Bipolar Disorder: Using Retrieval-Augmented Generation with an Open-Weight Large Language Model to Answer Clinical Questions Based on the CANMAT and ISBD 2018 Guidelines for Bipolar Disorder"

#### Supplementary Table 1: Example of a Clinical Vignette

| **Case 5, Section 3. Acute Management of Bipolar Mania** This is a 45-year-old man with a history of bipolar I disorder. He has had six prior hospitalizations for manic episodes, one with psychosis. He has no medical comorbidities. The patient presents with a four-week history of manic symptoms, including irritability, decreased need for sleep, pressured speech, and grandiose ideas. This was despite being on a therapeutic level of lithium, which he stopped taking a few days into mania. Current psychiatric medications include quetiapine 600 mg nightly, which has not helped despite him taking the medications under nurse supervision. Past psychiatric medications include oral paliperidone (not tolerated), and carbamazepine. The patient works irregularly as a contractor and lives alone.  You are wondering what medications should be provided for this patient’s mania. | |
| --- | --- |
| **Example of a rater’s query:**  What are the best medication options to acutely manage manic and possible psychotic symptoms. He relapsed on lithium and did not respond to add-on quetiapine 600 mg. Poor tolerability to paliperidone and carbamazepine. He is a 45 year old male. | |
| **Baseline LLM Response:** | **RAG LLM Response:** |
| for acute management of manic and psychotic symptoms in a patient with bipolar i disorder who has relapsed on **lithium** and did not respond to add-on **quetiapine**, consider:   - **olanzapine** as a first-line option due to its efficacy in treating manic episodes with psychotic features - **risperidone** as an alternative option, given its effectiveness in managing manic symptoms and psychosis - **asenapine** or **ziprasidone** as potential options, although they may be considered second-line due to varying efficacy and tolerability profiles - avoid **paliperidone** and **carbamazepine** due to poor tolerability - consider **valproate** as a potential mood stabilizer to augment antipsychotic therapy, if not already tried - monitor for side effects and adjust treatment as needed to optimize efficacy and tolerability | - given the patient has relapsed on **lithium** and did not respond to add-on **quetiapine** 600 mg, and has poor tolerability to **paliperidone** and **carbamazepine**, the following options can be considered: - **divalproex** as a monotherapy is a first-line option - **asenapine**, **aripiprazole**, **risperidone**, and **cariprazine** are also first-line monotherapy options - combination therapy with **aripiprazole + lithium/divalproex**, or **risperidone + lithium/divalproex**, or **asenapine + lithium/divalproex** can be considered, although **lithium** has already been tried - since the patient has possible psychotic symptoms, combination therapy may be preferred - considering the patient's poor tolerability to **paliperidone** and **carbamazepine**, **olanzapine**, **ziprasidone**, and **haloperidol** can be considered as second-line options, but their potential side effects and tolerability should be carefully evaluated. |

#### Supplementary Table 2. Prompts for baseline Llama model and RAG-based Llama model

| Baseline | Rag-based Model |
| --- | --- |
| ## Core Instructions  You are a clinical assistant helping physicians treat bipolar disorder. Answer in **bullet points** using markdown format.  - Only answer questions related to bipolar disorder.  - Keep your answer concise and accurate.  ## Patient-Specific Requirements  When a patient case is presented in the user query, you should identify:  1. **Disorder subtype**: Bipolar I or Bipolar II. If not specified, assume Bipolar I.  2. **Current mood state**: Depression, mania, hypomania, or euthymia/maintenance.  3. **Treatment line needed**: Determine first-line, second-line, or third-line.  4. **Patient Population**: Identify if the patient is of a specific population (e.g., pregnant, pediatric, elderly).  5. Identify whether the goal/intention of the query is to treat a certain episode, maintain, prevent relapse, or manage side effects etc.  ## Reasoning on Medication Selections  - Decide treatment recommendations based on subtype and mood state. Exclude failed medications.  - If patient has clinical features (eg. mixed features, anxiety, rapid cycling): Find treatment modifications for these features. Note preferred or avoided medications.  - If comorbidities exist (eg. diabetes, cardiovascular disease): Identify contraindicated medications. Exclude with reason.  - If the question specifies requirements (e.g., "medication without weight gain", "non-sedating"): Exclude medications that don't meet criteria.  - If a medication will cause a side effect, exclude any combination therapy containing it. Provide a short reason.  - If a certain mood episode needs to be prevented, then exclude medications that do not have data on preventing this phase. Prioritize medications with better evidence level.  ## Response Structure  - Provide first-line recommendations by default.  - If the first-line has failed, mention additional treatment lines.  - Use lowercase and **bold** for medication names.  - Use objective language (Do NOT say "I recommend...").  - No need to mention evidence level of the medication(eg. Level 1 or Level 2). | ## Core Instructions  You are a clinical assistant helping physicians treat bipolar disorder. Answer in **bullet points** using markdown format. Please follow these steps to answer the question:  1. Identify the key information needed to answer the question.  2. Locate relevant information in the context and IGNORE irrelevant information.  3. Extract or infer the answer from the provided context.  4. Provide your final answer.  ## Content Boundaries  - Remember: All your treatment/medication recommendations MUST be based on Guideline Contexts.  - Only answer questions related to bipolar disorder.  - Do not mention guideline source or tables in your responses.  - ONLY use information from the context.  - If the answer is NOT in the context, kindly respond that not enough information is available to answer the question.  - Keep your answer concise and accurate.  ## Patient-Specific Requirements  When a patient case is presented in the user query, you should identify:  1. **Disorder subtype**: Bipolar I or Bipolar II. If not specified, assume Bipolar I.  2. **Current mood state**: Depression, mania, hypomania, or euthymia/maintenance.  3. **Treatment line needed**: Determine first-line, second-line, or third-line.  4. **Patient Population**: Identify if the patient is of a specific population (e.g., pregnant, pediatric, elderly).  5. Identify whether the goal/intention of the query is to treat a certain episode, maintain, prevent relapse, or manage side effects etc.  ## Reasoning on Medication Selections  - Find the treatment table or recommendations based on subtype and mood state. Exclude failed medications.  - If patient has clinical features (eg. mixed features, anxiety, rapid cycling): Find treatment modifications for these features. Note preferred or avoided medications.  - If comorbidities exist (eg. diabetes, cardiovascular disease): Identify contraindicated medications. Exclude with reason.  - If the question specifies requirements (e.g., "medication without weight gain", "non-sedating"): Exclude medications that don't meet criteria.  - If a medication will cause a side effect, exclude any combination therapy containing it. Provide a short reason.  - If a certain mood episode needs to be prevented, then exclude medications that do not have data on preventing this phase. Prioritize medications with better evidence level.  ## Response Structure  - Provide first-line recommendations by default.  - If the first-line has failed, mention additional treatment lines.  - Use lowercase and **bold** for medication names.  - Use objective language (Do NOT say "I recommend...").  - No need to mention evidence level of the medication(eg. Level 1 or Level 2). |

#### Supplementary Table 3. Comparing correctness ratings for baseline and RAG models

|  | Total Answer Rated | |
| --- | --- | --- |
| Count | Baseline | RAG |
| Correct | 24 | 80 |
| Inaccurate | 54 | 36 |
| Wrong | 48 | 10 |
| Percentage |  |  |
| Correct | 19.05% | 63.49% |
| Inaccurate | 42.86% | 28.57% |
| Wrong | 38.10% | 7.94% |

#### Supplementary Figure 1: Confusion Matrix

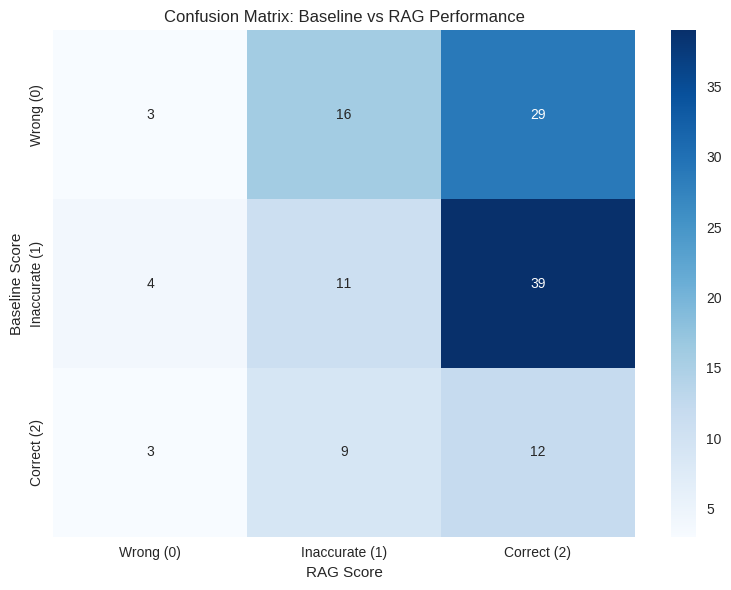

### Supplementary Document 1. Ordinal Mixed-Effects Model: Summary and Posterior Analysis

===== GENERAL ORDINAL MODEL RESULTS =====

Family: cumulative

Links: mu = logit

Formula: rating ~ method_binary + (1 | userLabel) + (1 | case)

Data: long_df (Number of observations: 252)

Draws: 4 chains, each with iter = 2000; warmup = 1000; thin = 1;

total post-warmup draws = 4000

Multilevel Hyperparameters:

~case (Number of levels: 21)

Estimate Est.Error l-95% CI u-95% CI Rhat Bulk_ESS Tail_ESS

sd(Intercept)	0.47	0.24	0.05	0.96 1.00	1080	1493

~userLabel (Number of levels: 6)

Estimate Est.Error l-95% CI u-95% CI Rhat Bulk_ESS Tail_ESS

sd(Intercept)	0.84	0.38	0.33	1.79 1.00	1473	2449

Regression Coefficients:

Estimate Est.Error l-95% CI u-95% CI Rhat Bulk_ESS Tail_ESS

Intercept[1]	-0.46	0.42	-1.28	0.42 1.00	1502	1903

Intercept[2]	1.68	0.44	0.84	2.63 1.00	1583	2084

method_binary	2.21	0.29	1.66	2.79 1.00	3217	2760

Further Distributional Parameters:

Estimate Est.Error l-95% CI u-95% CI Rhat Bulk_ESS Tail_ESS

disc	1.00	0.00	1.00	1.00	NA	NA	NA

Draws were sampled using sampling(NUTS). For each parameter, Bulk_ESS

and Tail_ESS are effective sample size measures, and Rhat is the potential

scale reduction factor on split chains (at convergence, Rhat = 1).

===== RATER DIFFERENCES (Mean Probability of 'Correct') =====

userLabel,0,1,diff_correct

participant1,0.17,0.61,0.44

participant2,0.16,0.6,0.44

participant4,0.2,0.66,0.46

participant5,0.08,0.41,0.33

participant6,0.29,0.77,0.47

participant7,0.26,0.73,0.47

===== RATER CREDIBLE INTERVALS =====

userLabel,method_binary,mean,lower,upper

participant1,0,0.17,0.09,0.27

participant1,1,0.61,0.47,0.75

participant2,0,0.16,0.09,0.25

participant2,1,0.6,0.45,0.73

participant4,0,0.2,0.12,0.3

participant4,1,0.66,0.53,0.78

participant5,0,0.08,0.04,0.14

participant5,1,0.41,0.26,0.57

participant6,0,0.29,0.18,0.44

participant6,1,0.77,0.64,0.87

participant7,0,0.26,0.15,0.39

participant7,1,0.73,0.6,0.84

===== RATER PROBABILITY OF SUPERIORITY (RAG > Baseline) =====

userLabel,prob_superiority

participant1,1

participant2,1

participant4,1

participant5,1

participant6,1

participant7,1
